# Normative Modeling of Structural and Diffusion MRI for Epilepsy Diagnosis and Classification

**DOI:** 10.64898/2026.11.14.26368080

**Authors:** Daniel J. Zhou, Marc Jaskir, Raina Hutchinson, Ethan S. Eisenberg, Julian Gal, Tamjid Imtiaz, Zack Goldblum, Chetan Vadali, Alfredo Lucas, Mariam Josyula, James J. Gugger, Erin C. Conrad, Sandhitsu Das, Joel M. Stein, Kathryn A. Davis, Nishant Sinha

## Abstract

**Background and Objectives:** Normative modeling expresses an individual’s brain measures as deviations from a large healthy reference and has been advanced as a route to precision imaging biomarkers in epilepsy, where structural abnormality is distributed rather than confined to the presumed focus. Whether this framework can inform clinical classification from clinically acquired T1-weighted and diffusion MRI is untested. We asked whether normative deviations discriminate between (1) epilepsy and functional seizures; (2) focal and generalized epilepsy; and (3) drug-resistant and drug-responsive epilepsy.

**Methods:** We retrospectively studied patients with epilepsy or functional seizures without comorbid epilepsy who underwent 3-Tesla epilepsy-protocol MRI at the University of Pennsylvania. Morphometric deviation scores (cortical thickness, subcortical volume) were derived from CentileBrain (*n*>37,000 healthy references). White matter fractional anisotropy and mean diffusivity profiles were referenced to harmonized Human Connectome Project cohorts. Lobar composites, hemispheric asymmetry indices, and whole-tract means were compared with rank-sum tests; discrimination was summarized as area under the ROC curve (AUC) with bootstrap confidence intervals and Benjamini-Hochberg correction. Whole-brain deviation burden was examined as a secondary analysis. Minimum detectable effects were estimated by Monte Carlo simulation at 80% power (α=0.05).

**Results:** We included 273 patients with epilepsy (203 focal, 37 generalized, 33 unclassified; 97 drug-resistant, 176 drug-responsive) and 32 with functional seizures. In the primary analyses, no regional or tract-level normative measure distinguished epilepsy from functional seizures, focal from generalized epilepsy, or drug-resistant from drug-responsive epilepsy; AUCs did not exceed 0.62, with upper confidence limits up to 0.72. In the secondary analysis, whole-brain morphometric deviation burden was modestly higher in epilepsy than in functional seizures (AUC 0.66, 95% CI 0.57-0.74) but did not separate the within-epilepsy comparisons. The study could detect AUC ≥0.65 for epilepsy versus functional seizures, ≥0.64 for focal versus generalized, and ≥0.60 for drug-resistant versus drug-responsive epilepsy.

**Discussion:** Univariate normative deviation summaries from clinical MRI do not currently distinguish these groups well enough to guide care. Whole-brain deviation burden, the only summary measure that differed after correction, was detectable at the group level but insufficient for individual-level classification. Progress may require network-targeted analysis of patient-specific deviations and multimodal integration.

## Introduction

The evaluation and management of patients with suspected epilepsy involves a sequence of clinical decisions, each carrying diagnostic or prognostic uncertainty. The first is determination of whether the paroxysmal events are epileptic in origin. Functional seizures, also known as psychogenic nonepileptic seizures or events, are common, frequently mistaken for epilepsy, and a leading cause of misdiagnosis, delayed care, and unnecessary antiseizure medication (ASM) exposure.^1,2^ The second is classification of epilepsy as focal or generalized, which differ in presumed mechanism, treatment, and surgical candidacy. The third is identifying ASM resistance, as approximately one-third of patients are drug-resistant, defined by the International League Against Epilepsy as failure of two adequate trials of appropriately chosen and tolerated antiseizure medications to achieve sustained seizure freedom.^3,4^ While growing evidence suggests that earlier surgical treatment in patients with drug-resistant epilepsy improves seizure freedom outcomes, the clinical determination of drug resistance depends on sequential trials of ASMs and is reached at a median of 10 years after epilepsy diagnosis.^5–7^ Objective biomarkers could inform any of these decisions, enabling earlier interventions and improved outcomes.

Quantitative analysis of brain MRI offers objective measures that may be relevant to each of these clinical classifications. From T1-weighted images, cortical thickness and subcortical volumes capture morphometric change, while diffusion-weighted imaging yields measures of white matter microstructure such as fractional anisotropy (FA) and mean diffusivity (MD).^8,9^ In epilepsy, structural abnormalities have been found to be widespread, extending beyond the presumed seizure focus, including distributed cortical thinning and subcortical volume loss, and altered white matter microstructure most pronounced in the corpus callosum, cingulum, and external capsule.^10,11^ Compared to generalized epilepsy, focal epilepsy showed more extensive white matter abnormalities and partly distinct regional patterns of gray matter change, alongside differences in structural network organization.^10,12,13^ In functional seizures, the few studies comparing patients to healthy controls reported subtle, network-level differences. These included convergent cortical thinning in visual and sensorimotor cortex and hyperconnectivity across sensorimotor, default-mode, and attentional networks, though findings came from small, heterogeneous cohorts.^14,15^ Few studies have directly compared imaging between drug-resistant and drug-responsive epilepsy. In idiopathic generalized epilepsy, drug-resistant patients showed more widespread FA alterations in white matter networks.^16^ Structural network analyses in temporal lobe epilepsy revealed differences in global network architecture between the two groups.^17,18^

A persistent challenge across these comparisons is the heterogeneity of epilepsy, which limits the interpretability of traditional group-level analyses. Case-control comparisons rely on population averages and may obscure meaningful individual-level variation in brain structure, a limitation compounded in focal epilepsy by uncertainty in epileptogenic network localization, particularly in cases without invasive monitoring or surgery. Normative modeling addresses this limitation by quantifying how much an individual deviates from a reference distribution matched by age and sex, rather than relying on group averages, yielding interpretable individual-level deviation scores.^19^ This framework has recently been advanced as a route to precision MRI biomarkers in epilepsy, with potential for abnormality localization and individual-level prediction.^20^

Here we ask whether normative modeling is ready to inform the clinical classifications providers rely on, using clinically acquired MRI, which at our tertiary epilepsy center includes 1-mm T1-weighted and 30-direction diffusion-weighted imaging. We applied normative models of cortical thickness, subcortical volume, and along-tract white matter microstructure to the scans from a consecutive tertiary-center cohort, and tested whether individual-level deviations can discriminate between epilepsy and functional seizures, between focal and generalized epilepsy, and between drug-resistant and drug-responsive epilepsy. Rather than asking whether group differences exist, we asked whether individual-level deviations discriminate these groups strongly enough to be clinically useful. We also estimated the effect sizes this cohort could detect and the range of effects compatible with the data, so that negative findings could be interpreted as bounds.

## Materials and methods

### Patient Selection

This retrospective study was approved by the Institutional Review Board of the University of Pennsylvania (protocol 835008) with a waiver of informed consent. Normative reference data were obtained from the Human Connectome Project Young Adult and Aging datasets under their respective data use agreements; participants in those studies provided written informed consent at the originating institutions.

Epilepsy-protocol 3-Tesla MRI brain scans with both T1-weighted and 30-direction diffusion-weighted sequences were obtained from patients who were seen at the Penn Epilepsy Center at the University of Pennsylvania from May 2022 to January 2025. Medical records were reviewed by a board-certified epileptologist (D.Z.) to determine each patient’s clinical diagnosis and classification, based on the assessments of their treating epileptologists.

Patients with epilepsy and patients with functional seizures without comorbid epilepsy were included. Patients without a clear diagnosis of either, including those with functional seizures occurring alongside epilepsy, single unprovoked or acute symptomatic seizures, or events that were not clearly epileptic, were excluded. Among patients with epilepsy, those with prior brain surgery before the time of MRI, or without usable diffusion or morphometric data, were excluded.

Epilepsy was classified as focal, generalized, or unclassified according to the treating epileptologist’s diagnosis. Among patients treated with ASM for at least one year, drug resistance was defined according to the International League Against Epilepsy as failure of two tolerated, appropriately chosen and used ASM schedules to achieve sustained seizure freedom,^4^ assessed at the most recent follow-up. Patients were classified as drug-responsive if they had been seizure-free on ASM for at least 12 consecutive months, with seizure freedom continuing through the most recent follow-up. Patients whose treatment response could not be clearly determined, such as those with continued seizures after only a single medication trial, were excluded from the study (Supplementary Fig. 1).

Extracted clinical metadata included age at first seizure, epilepsy duration, history of convulsive seizures or status epilepticus, presence of an epileptogenic lesion on MRI, and presence of interictal epileptiform discharges on routine or ambulatory EEG. Epilepsy duration was calculated from the date of first seizure to the date of the MRI scan. Presence of an epileptogenic lesion was determined by the treating epileptologist.

### Image Acquisition and Processing

MRI scans of the clinical cohort were acquired at the Hospital of the University of Pennsylvania on Siemens 3T scanners. T1-weighted images were acquired using a magnetization-prepared rapid gradient-echo (MPRAGE) sequence at 1.0 mm isotropic resolution. Diffusion-weighted images were acquired with a single-shell echo-planar imaging sequence comprising 30 diffusion-encoding directions at b = 1000 s/mm² and one non-diffusion-weighted (b = 0) volume, at a native resolution of 2.2 mm isotropic. A separate b = 0 volume with reversed phase-encoding direction was acquired to estimate the susceptibility-induced B0 field, and the resulting geometric distortions were corrected during preprocessing using FSL topup.^21^

Cortical thickness and subcortical volumes were estimated from T1-weighted images using FreeSurfer (version 7.3.2)^22^ with the Desikan-Killiany atlas,^23^ yielding 68 cortical and 14 subcortical regions of interest. Diffusion-weighted images were preprocessed using QSIPrep (version 1.0.1)^24^ and resampled to 1.25 mm isotropic resolution. White matter tract profiles were reconstructed using pyAFQ (version 2.1),^25^ yielding FA and MD, sampled at 100 nodes along each of 16 bundles. The middle cerebellar peduncle was excluded, as brainstem tractography is prone to susceptibility-induced distortion and artifactual contamination by crossing fibers.^26^

### Normative Modeling and Data Harmonization

Deviation scores were derived separately for morphometry and tractometry, each expressing an individual’s brain measures as a deviation from an age- and sex-matched healthy reference population (Fig. 1).

**Figure 1.**
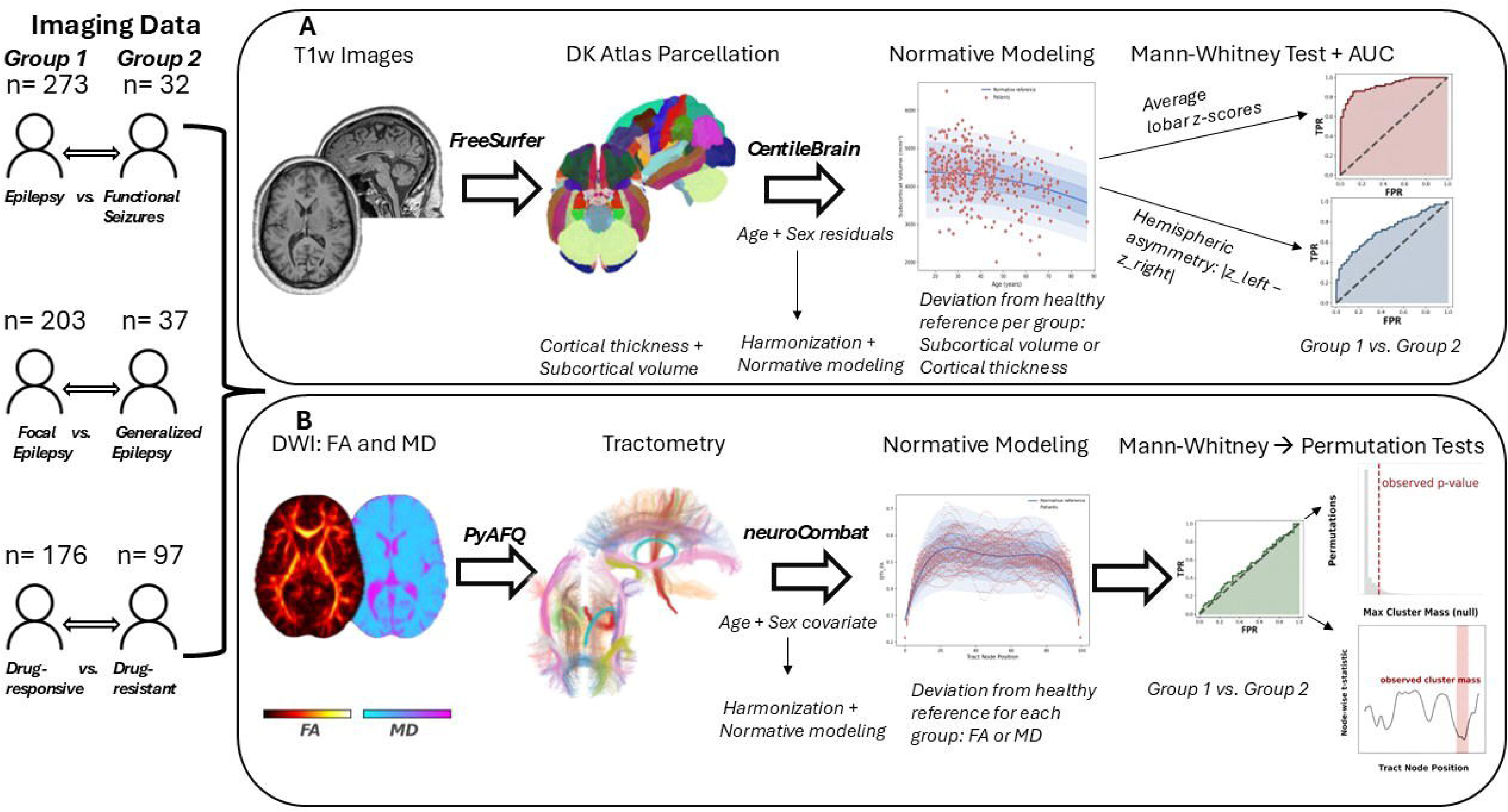
Study design and analysis pipeline. The left column shows the three comparisons: epilepsy vs. functional seizures, focal vs. generalized epilepsy, and drug-resistant vs. drug-responsive epilepsy. (A) Morphometry: T1-weighted images were parcellated with FreeSurfer (Desikan-Killiany atlas), and regional cortical thickness and subcortical volume were converted to deviation z-scores with CentileBrain. (B) Tractometry: fractional anisotropy (FA) and mean diffusivity (MD) profiles from pyAFQ were harmonized with neuroCombat and converted to z-scores against Human Connectome Project references. The normative-model panels show example study data; other plots are schematic.

For morphometry, z-scores for cortical thickness and subcortical volume were obtained from CentileBrain, which used pretrained sex-specific models expressing each region as a standardized residual adjusting for age, sex, intracranial volume for subcortical volumes, and mean cortical thickness for cortical thickness.^27^ These models were derived from over 37,000 healthy individuals aged 3-90. Cortical thickness and subcortical volumes were extracted with FreeSurfer 7.3.2, and the corresponding scanner vendor (Siemens) and FreeSurfer version were provided to CentileBrain as covariates.

For tractometry, z-scores for FA and MD along tracts were referenced to healthy subjects from the Human Connectome Project (HCP) Young Adult (ages 22-35, 1064 subjects) and Aging (ages 36-100+, 1365 subjects) datasets,^28,29^ preprocessed with the HCP minimal preprocessing pipelines.^30^ Diffusion tensors were fit to the b = 1000 s/mm² shell for HCP Young Adult and to the b = 1500 s/mm² shell for HCP Aging, which has no b = 1000 shell. Missing left-fornix profiles (22 clinical and 46 HCP scans) were replaced with the pooled mean. Site- and protocol-related variance was removed with neuroCombat (version 0.2.12),^31^ with acquisition site as the batch variable (six sites: HCP Young Adult, the four HCP Aging sites, and Penn), age and sex as protected covariates, and site parameters estimated from all scans without a reference batch. As deviations shared by all patients may be partly absorbed into site effects and harmonization can distort the normative age curve, we repeated all tractometry comparisons without harmonization and with site parameters estimated from reference subjects only (Supplementary Fig. 4). Following harmonization, each tract and metric was regressed on age and sex in the reference subjects only (the HCP cohorts and 26 syncope patients who underwent the same epilepsy-protocol imaging), and each patient’s value was expressed as a standardized residual. Deviations for the 42 patients (14%) younger than 22 years of age (39 with epilepsy, 3 with functional seizures) relied on extrapolation below the HCP age range. The syncope reference patients had a median age of 33.4 years (IQR 26.0-48.6), versus 36.0 years (IQR 26.0-46.0) in the analyzed cohort.

Region-level measures were aggregated for statistical analyses. For morphometry, ROI z-scores were averaged (unweighted) within seven regional groups (frontal, temporal, parietal, occipital, insula, thalamus, and basal ganglia), combining cortical and subcortical structures where both contributed (hippocampus and amygdala within the temporal group; caudate, putamen, pallidum, and nucleus accumbens as basal ganglia; thalamus alone). Composite z-scores retained their sign, so that negative values indicate lower thickness or volume than expected. A lobe-level asymmetry index was computed as the absolute difference between left and right hemisphere mean z-scores, without normalization by the bilateral sum, which is unstable when z-scores approach zero. For tractometry, left and right homologues of each paired tract were averaged node-by-node, and commissural tracts were used as reconstructed, yielding nine tract profiles (six paired bilateral tracts and three commissural tracts) of 100 nodes each. For each profile, a tract-level mean z-score summarized overall deviation, while the full node-wise profile was retained for along-tract analysis.

### Statistical Analyses

Analyses were performed in Python using scipy and statsmodels.^32,33^ Group demographics were compared using Fisher’s exact test for categorical variables and Mann-Whitney U tests for continuous variables. Three imaging comparisons were performed: epilepsy versus functional seizures, focal versus generalized epilepsy, and drug-resistant versus drug-responsive epilepsy. Because normative z-scores were derived from age- and sex-stratified reference models, no additional covariate adjustment was applied in the primary analyses.

Imaging features were analyzed at two scales: region/tract-level summary scores and node-wise along-tract profiles. Lobar composite z-scores, hemispheric asymmetry indices (|z_left_ − z_right_|), and tract-level mean z-scores (FA and MD) were compared using Mann-Whitney U tests. Discriminability was quantified as the area under the curve (AUC = P(group_a > group_b)), reported as max(AUC, 1 − AUC) with the direction of each effect given separately, with 95% confidence intervals from 5,000 bootstrap resamples. Benjamini-Hochberg correction was applied within each feature family per comparison (lobar composite z-scores, asymmetry indices, and pooled FA+MD tract z-scores). Two secondary analyses were performed. First, whole-brain deviation burden was summarized per patient as the number of regions with |*z*|>2 and the mean |*z*| across the 82 morphometric regions, the 18 tract-level FA and MD z-scores, and the 3,000 node-level z-scores of the 15 bundles without left-right averaging, with correction across these six measures; the epilepsy versus functional seizures comparison was repeated in non-lesional epilepsy. Second, the mean hippocampal z-score and hippocampal asymmetry index were compared, with correction across the two measures. As a positive control, these measures were compared between patients with unilateral mesial temporal sclerosis and no other lesion and patients with non-lesional epilepsy, and the sclerotic hippocampus was compared with the contralateral side (Wilcoxon signed-rank test).

Along-tract differences in FA and MD were assessed using cluster-based permutation testing, without covariate adjustment. Group labels were randomly permuted 5,000 times; for each permutation, a t-statistic was computed at each node comparing groups, contiguous suprathreshold nodes (*p*<0.05) were grouped into clusters, and the maximum cluster mass was retained to form a null distribution specific to that tract. Observed clusters were tested against this tract-specific null, controlling the family-wise error rate across nodes within each tract. As all bilateral tracts were surveyed, along-tract findings were considered exploratory.

Minimum detectable effect sizes were estimated for each comparison by Monte Carlo simulation (3,000 iterations per candidate effect size), assuming normally distributed scores with equal variance and a location shift between groups, using each comparison’s achieved sample sizes, 80% power, and a two-sided Mann-Whitney U test at α=0.05, both uncorrected and at α=0.05/18 (a single true effect within the largest feature family after correction). Effects were expressed as the minimum detectable AUC.

### Data Availability

De-identified derived data supporting the findings of this study, including regional morphometric and tractometric z-scores with accompanying clinical classifications, will be made publicly available in PennSieve upon publication.^34^ Analysis code is available at https://github.com/penn-cnt/Epilepsy-Normative-MRI-Release. Normative reference data are available through ConnectomeDB (HCP Young Adult) and the NIMH Data Archive (HCP Aging), and CentileBrain models are available at centilebrain.org. Raw imaging data may be shared with qualified investigators under a data use agreement, subject to institutional approval.

### AI Use

The authors used Claude Opus 5.5 (Anthropic, 2026) to improve the clarity and readability of the manuscript and for coding assistance in developing the Python analysis code. The scientific content, interpretation, and conclusions were written entirely by the authors, and all AI-suggested revisions and code were reviewed and approved by the authors.

## Results

### Demographic and clinical information

Demographic and clinical characteristics are summarized in Table 1. We identified 781 patients who underwent a clinical 3-Tesla epilepsy-protocol brain MRI at the University of Pennsylvania. After exclusions, the analyzed cohort comprised 273 patients with epilepsy and 32 with functional seizures without comorbid epilepsy (Supplementary Fig. 1). Within the epilepsy cohort, 203 (74%) had focal epilepsy, 37 (14%) generalized, and 33 (12%) unclassified; 176 (64%) were drug-responsive and 97 (36%) drug-resistant. Patients with epilepsy and functional seizures did not differ in age at MRI but differed in sex distribution (*p*<0.001; Table 1). Focal and generalized epilepsy differed in age at onset, age at MRI, history of convulsive seizures, and presence of an epileptogenic lesion (all *p*<0.05), but not in sex or status epilepticus history. Drug-responsive and drug-resistant patients differed in age at onset, epilepsy duration, and history of convulsive seizures (all *p*<0.05).

**Table 1.**
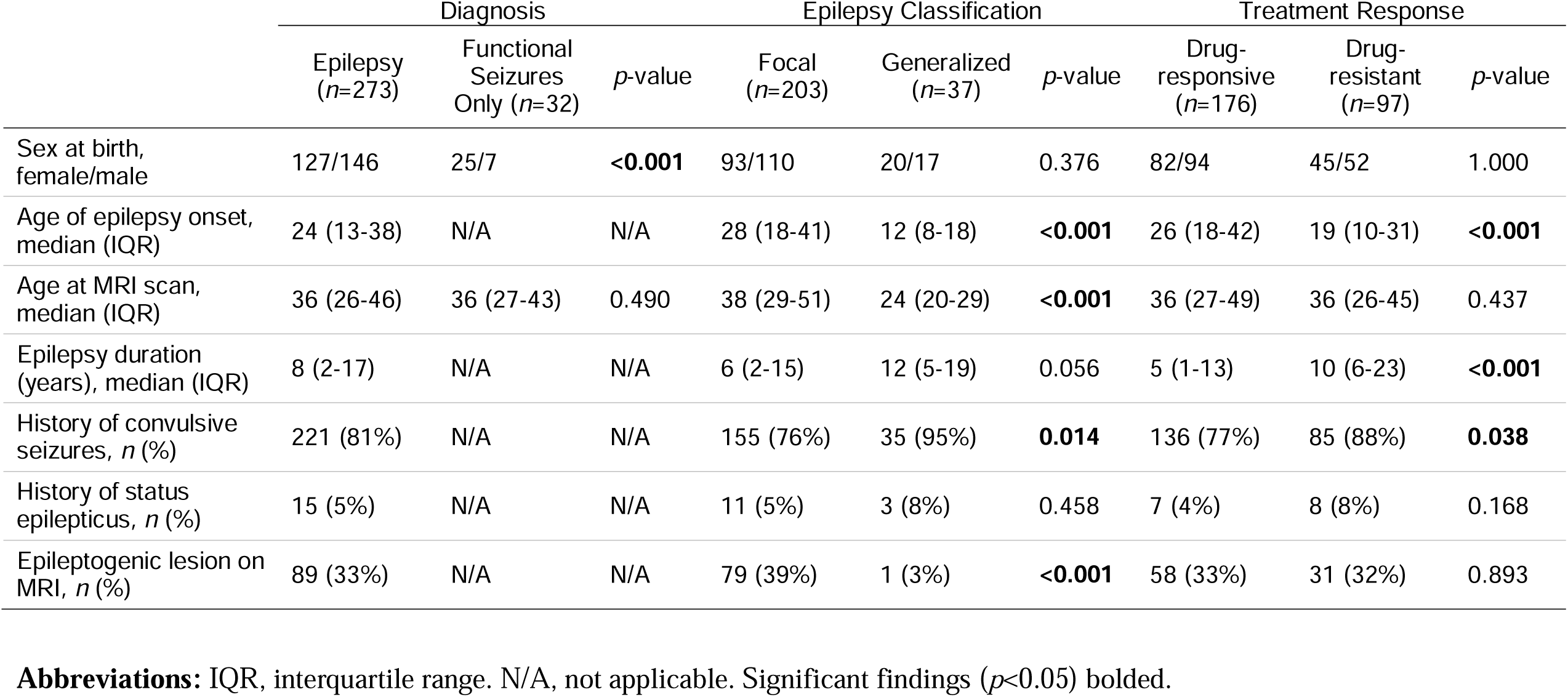
Demographic and clinical variables compared between groups.

|  | Diagnosis |  |  | Epilepsy Classification |  |  | Treatment Response |  |  |
|---|---|---|---|---|---|---|---|---|---|
|  | Epilepsy (*n*=273) | Functional Seizures Only (*n*=32) | *p*-value | Focal (*n*=203) | Generalized (*n*=37) | *p*-value | Drug-responsive (*n*=176) | Drug-resistant (*n*=97) | *p*-value |
| Sex at birth, female/male | 127/146 | 25/7 | <**0.001** | 93/110 | 20/17 | 0.376 | 82/94 | 45/52 | 1.000 |
| Age of epilepsy onset, median (IQR) | 24 (13-38) | N/A | N/A | 28 (18-41) | 12 (8-18) | <**0.001** | 26 (18-42) | 19 (10-31) | <**0.001** |
| Age at MRI scan, median (IQR) | 36 (26-46) | 36 (27-43) | 0.490 | 38 (29-51) | 24 (20-29) | <**0.001** | 36 (27-49) | 36 (26-45) | 0.437 |
| Epilepsy duration (years), median (IQR) | 8 (2-17) | N/A | N/A | 6 (2-15) | 12 (5-19) | 0.056 | 5 (1-13) | 10 (6-23) | <**0.001** |
| History of convulsive seizures, *n* (%) | 221 (81%) | N/A | N/A | 155 (76%) | 35 (95%) | **0.014** | 136 (77%) | 85 (88%) | **0.038** |
| History of status epilepticus, *n* (%) | 15 (5%) | N/A | N/A | 11 (5%) | 3 (8%) | 0.458 | 7 (4%) | 8 (8%) | 0.168 |
| Epileptogenic lesion on MRI, *n* (%) | 89 (33%) | N/A | N/A | 79 (39%) | 1 (3%) | <**0.001** | 58 (33%) | 31 (32%) | 0.893 |

**Abbreviations:** IQR, interquartile range. N/A, not applicable. Significant findings (*p*<0.05) bolded.

### Epilepsy versus functional seizures

Lobar composite and asymmetry z-scores did not differ significantly between epilepsy and functional seizures after FDR correction (Fig. 2A-B). In tractometry, no tract differed at the whole-tract level (Fig. 2C-D), and along-tract analysis showed no significant clusters. These results held without harmonization and with reference-estimated site parameters estimated from reference subjects only (Supplementary Fig. 4).

**Figure 2.**
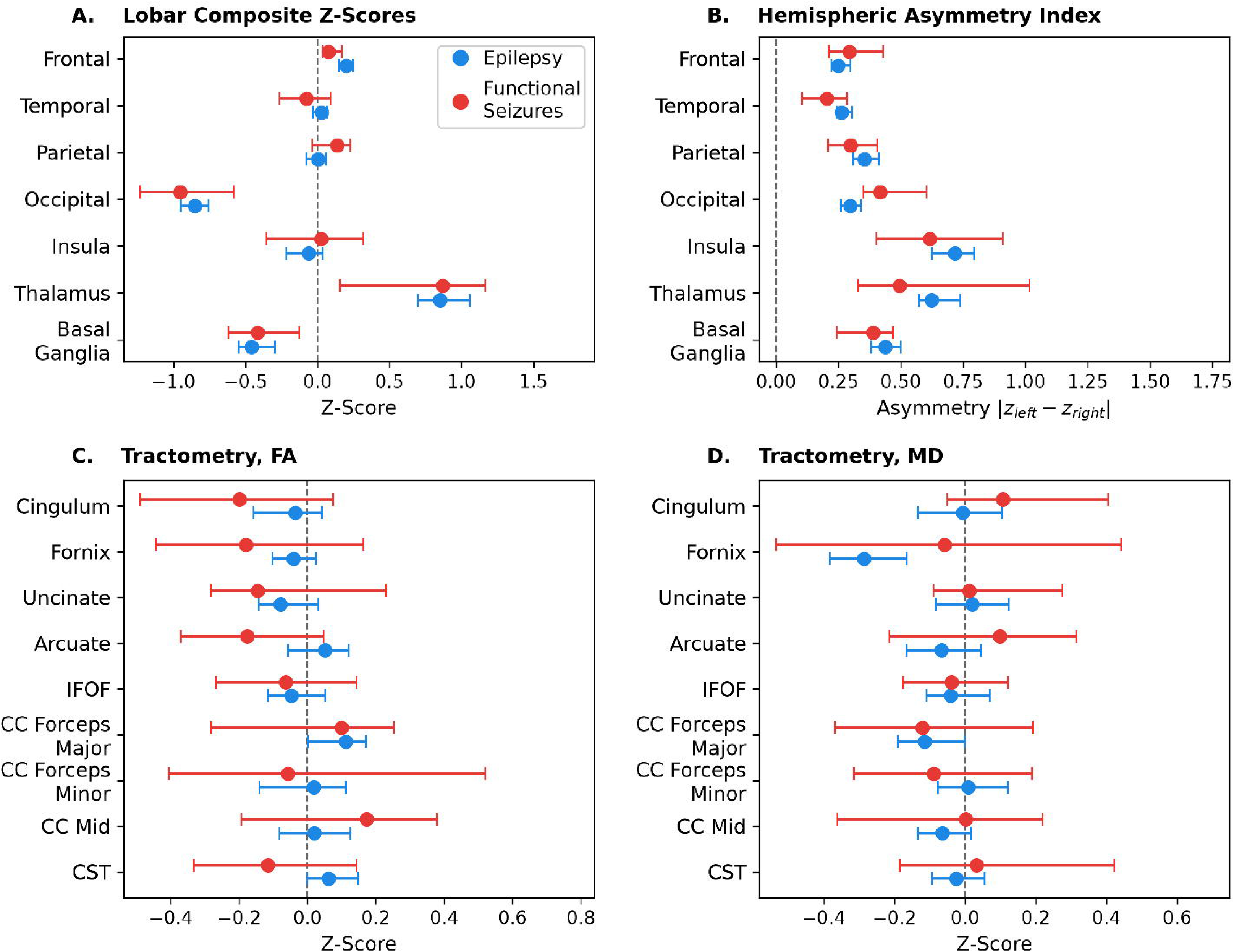
Normative deviations in epilepsy versus functional seizures. (A) Lobe-level composite z-scores and (B) hemispheric asymmetry index across seven regions. (C-D) Bilateral tract-averaged z-scores for fractional anisotropy (FA) and mean diffusivity (MD) across nine white matter tracts. Z-scores reflect deviation from a normative reference population, with negative values indicating lower regional volume or cortical thickness (A) or lower FA or MD (C-D). The asymmetry index (B) represents the absolute difference between left and right hemisphere z-scores within each region. Dots show group medians, and bars show bootstrap 95% confidence intervals of the median (5,000 resamples). Dashed vertical lines indicate the normative reference (*z*=0) or perfect hemispheric symmetry. No difference reached significance after FDR correction.

### Focal versus generalized epilepsy

Lobe-level composite and asymmetry z-scores did not differ significantly between focal and generalized epilepsy after FDR correction (Fig. 3A-B). In tractometry, no tract differed significantly at the whole-tract level (Fig. 3C-D). Without harmonization, eight MD tracts separated the groups (AUC up to 0.72, q<0.05), but this reflected a miscalibrated reference age curve, as the b-value difference between HCP cohorts shifted the median MD z-score of reference subjects aged 36-50 to about -1.0, and focal and generalized epilepsy differed in age (Supplementary Fig. 4). No tract differed when age and sex were protected and site parameters were estimated from reference subjects only. Exploratory along-tract analysis revealed several differences: compared with generalized epilepsy, focal epilepsy showed lower FA in the central segment of the forceps minor (*p*=0.007), and higher MD in the mid-segment and posterior segments of the inferior fronto-occipital fasciculus (*p*=0.016 and 0.043, respectively), posterior arcuate fasciculus (*p*=0.022), and anterior forceps major (*p*=0.038) (Supplementary Fig. 2).

**Figure 3.**
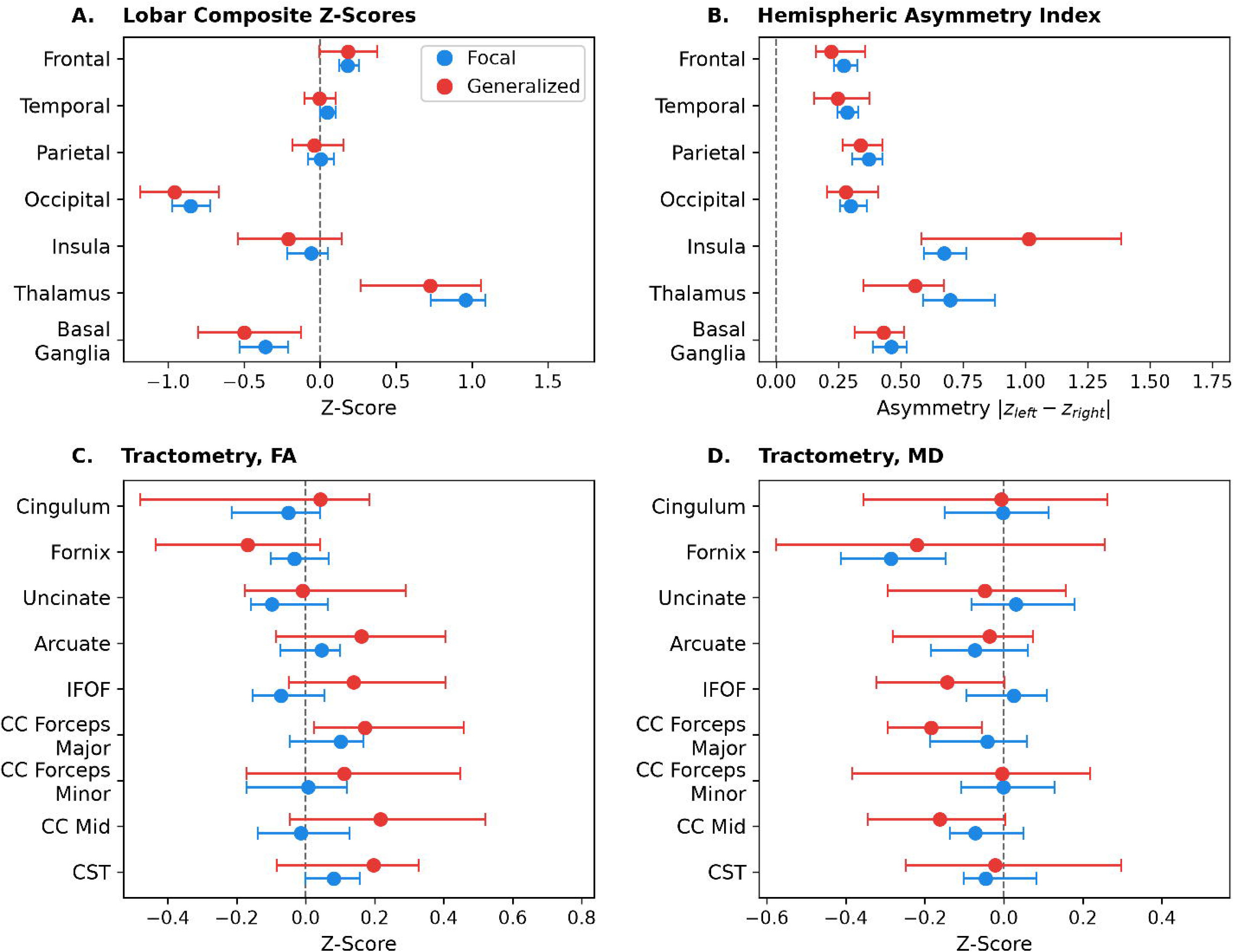
Normative deviations in focal versus generalized epilepsy. (A) Lobe-level composite z-scores and (B) hemispheric asymmetry index across seven regions. (C-D) Bilateral tract-averaged z-scores for fractional anisotropy (FA) and mean diffusivity (MD) across nine white matter tracts. Dots show group medians, and bars show bootstrap 95% confidence intervals of the median (5,000 resamples). Dashed vertical lines indicate the normative reference (*z*=0) or perfect hemispheric symmetry. No difference reached significance after FDR correction.

### Drug-responsive versus drug-resistant epilepsy

Lobe-level composite and asymmetry z-scores did not differ significantly between drug-responsive and drug-resistant epilepsy after FDR correction (Fig. 4A-B). In tractometry, no tract differed significantly at the whole-tract level after FDR correction (Fig. 4C-D). Exploratory along-tract analysis localized higher MD in drug-resistant epilepsy to the anterior cingulum (*p*=0.017), and lower FA in drug-resistant epilepsy to the mid-segment of the inferior fronto-occipital fasciculus (*p*=0.048) (Supplementary Fig. 3). In a sensitivity analysis with age and sex protected and site parameters estimated from reference subjects only, three tract differences reached significance at AUC ≈0.60 (cingulum MD, arcuate fasciculus FA, and inferior fronto-occipital fasciculus FA; Supplementary Fig. 4).

**Figure 4.**
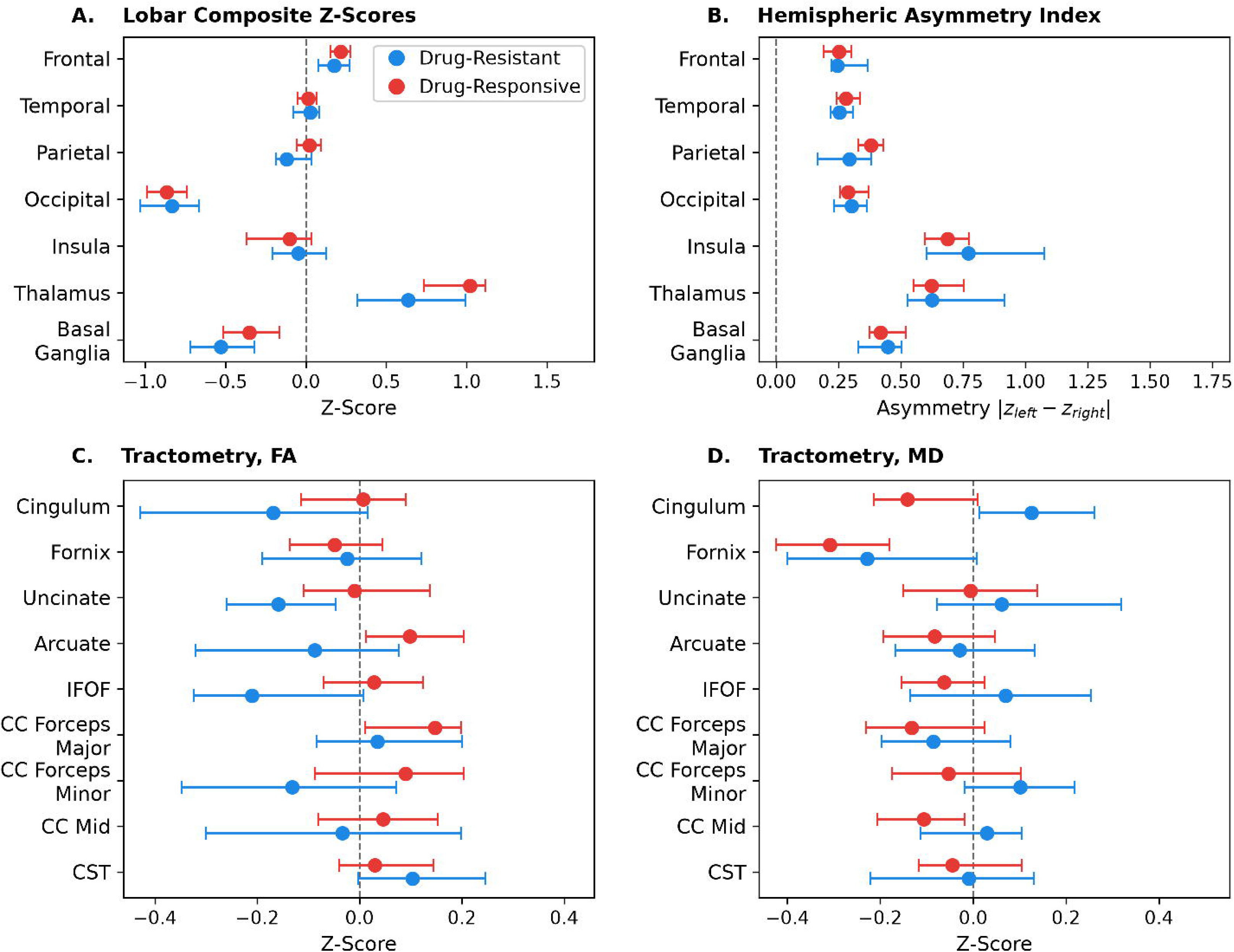
Normative deviations in drug-resistant versus drug-responsive epilepsy. (A) Lobe-level composite z-scores and (B) hemispheric asymmetry index across seven regions. (C-D) Bilateral tract-averaged z-scores for fractional anisotropy (FA) and mean diffusivity (MD) across nine white matter tracts. Dots show group medians, and bars show bootstrap 95% confidence intervals of the median (5,000 resamples). Dashed vertical lines indicate the normative reference (*z*=0) or perfect hemispheric symmetry. No difference reached significance after FDR correction.

### Secondary analyses

Whole-brain morphometric deviation burden was modestly higher in epilepsy than in functional seizures (median 7 vs 5 of 82 regions with |*z*|>2; AUC 0.66, 95% CI 0.57-0.74, *q*=0.010; mean |z| AUC 0.67, *q*=0.009; Supplementary Fig. 5A). When restricted to patients with non-lesional epilepsy, the difference from functional seizures was attenuated and did not survive correction (*n*=184 vs 32; regions with |*z*|>2 AUC 0.62, 95% CI 0.53-0.72, *q*=0.08; mean |*z*| AUC 0.64, 95% CI 0.54-0.74, *q*=0.07), and patients with lesional epilepsy had the highest burden (median 9 regions of interest). Burden did not differ between focal and generalized or drug-resistant and drug-responsive epilepsy, and tractometric burden did not differ in any comparison (Supplementary Table 1).

Hippocampal measures (mean z-score and asymmetry) did not discriminate any of the three comparisons (Supplementary Table 1). Among 12 patients with unilateral mesial temporal sclerosis and no other lesion (6 left, 6 right), the sclerotic hippocampus was smaller than the contralateral side (median difference -1.00; Wilcoxon *p*=0.007), by ≥3 standard deviations in 4 of 12 (Supplementary Fig. 5B). Hippocampal asymmetry in these patients was correspondingly greater than in non-lesional epilepsy (median 1.00 vs 0.48; AUC 0.72, 95% CI 0.55-0.88, *q*=0.02; Supplementary Fig. 5C), whereas the mean hippocampal z-score did not differ significantly.

### Minimum detectable effect sizes

At 80% power and uncorrected α=0.05, the study could detect AUC ≥0.65 for epilepsy versus functional seizures, ≥0.64 for focal versus generalized, and ≥0.60 for drug-resistant versus drug-responsive epilepsy (Table 2). For a single true effect within the largest correction family (18 tract features), these increased to 0.70, 0.69, and 0.64. The largest observed AUCs were 0.61 (95% CI 0.50-0.72), 0.62 (0.53-0.70), and 0.60 (0.53-0.67), respectively (Fig. 5; Supplementary Table 1).

**Figure 5.**
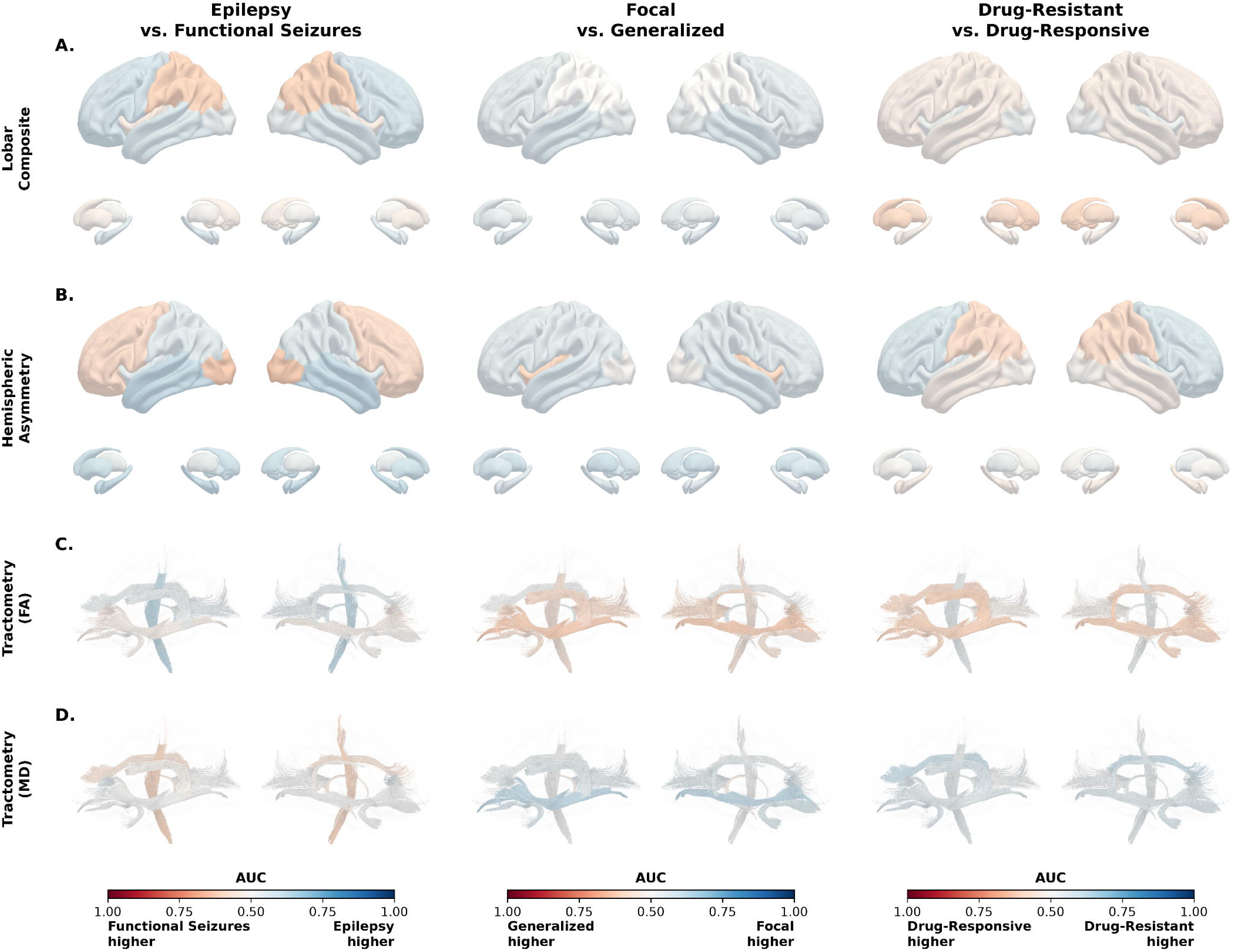
Discrimination of normative deviation features across comparisons. Columns show epilepsy vs functional seizures, focal vs generalized epilepsy, and drug-resistant vs drug-responsive epilepsy. (A) Lobar composite z-scores, shown on the lateral cortical surface (top) and on subcortical structures in lateral and medial views (bottom). (B) Hemispheric asymmetry index. (C) Whole-tract mean fractional anisotropy (FA). (D) Whole-tract mean diffusivity (MD). Each region or tract is colored by max(AUC, 1 − AUC) on a 0.50-1.00 scale, with hue indicating the group with higher values. Hippocampus and amygdala are colored by the temporal composite; no feature reached significance after Benjamini-Hochberg correction (q<0.05).

**Table 2.** Minimum detectable effect sizes for each comparison.

| Comparison | n<br>(group 1) | n<br>(group 2) | MDE AUC<br>(uncorrected) | MDE AUC<br>(corrected) |
|---|---|---|---|---|
| Epilepsy vs Functional Seizures | 273 | 32 | 0.65 | 0.70 |
| Focal vs Generalized | 203 | 37 | 0.64 | 0.69 |
| Drug-Resistant vs Drug-Responsive | 97 | 176 | 0.60 | 0.64 |

**Abbreviations:** AUC, area under the receiver operating characteristic curve; MDE, minimum detectable effect. *n* values are given per group in the order named in each comparison. MDEs are the smallest AUC detectable with 80% power under a two-sample normal-shift model, at uncorrected α=0.05 and at α=0.05/18, the Benjamini-Hochberg threshold for a single true effect in the largest correction family (18 tract features).

## Discussion

Normative modeling of clinical epilepsy-protocol MRI did not reliably discriminate between the clinical groups we tested. In the primary analyses, no regional or tract-level measure (lobar composite, hemispheric asymmetry, or whole-tract mean) distinguished epilepsy from functional seizures, focal from generalized epilepsy, or drug-resistant from drug-responsive epilepsy. In secondary analyses, neither hippocampal measure separated the groups, and the one significant difference was in whole-brain deviation burden, which was higher in epilepsy than in functional seizures (AUC 0.66) and partly reflected visible lesions (AUC 0.62 in non-lesional epilepsy), an effect detectable at the group level but insufficient for individual-level classification. Even the largest regional or tract-level effect compatible with these data (upper confidence limit 0.72) would not be actionable. Under a binormal model, an AUC of 0.72 corresponds to about 32% sensitivity at 90% specificity, whereas discrimination conventionally considered outstanding (AUC ≥0.90)^35^ would be a reasonable minimum before such a marker could inform decisions such as withdrawing ASM. Within the precision of this cohort, univariate normative deviation summaries did not reach a level of discrimination that could inform clinical care.

The gap between group-level effects and individual-level discrimination has also been reported for normative modeling in other disorders. In schizophrenia, normative modeling of harmonized diffusion MRI in 601 patients and 512 controls produced larger group-level effects in deviation scores than in raw measures, yet summary deviation measures classified individuals at AUC below 0.63; only combining tracts and diffusion measures exceeded 0.70.^36^ Our findings followed the same pattern: whole-brain deviation burden and hippocampal asymmetry differed at the group level, but neither discriminated individuals well. In temporal lobe epilepsy, supervised classification of multimodal normative deviations reached AUC 0.77 for distinguishing patients from disease controls and 0.64 for predicting postsurgical seizure freedom.^37^ Together, these studies indicate that combining deviations across regions and modalities could improve on single summary scores, but that discrimination with multimodal models remains below the level required for clinical decision-making.

Structural correlates have been reported in both epilepsy and functional seizures, but they differ in extent and consistency. In functional seizures, findings have been subtle and variable across studies, including superior temporal and occipital cortical changes and reduced motor and premotor cortical thickness and volume.^38,39^ In epilepsy, large-scale analyses have established widespread cortical thinning and subcortical volume reduction, but the abnormalities shared by all epilepsy groups are modest against healthy controls: lower right thalamic volume (Cohen’s d 0.24-0.73) and lower bilateral precentral cortical thickness (d 0.34-0.52), corresponding to AUCs of roughly 0.57-0.70 and 0.60-0.64.^10^ These effects lie largely at or below this cohort’s minimum detectable effect, and the expected separation from functional seizures, which carry their own structural deviations, is smaller than that from healthy controls, consistent with the weak separation observed here.

The hippocampal positive control illustrates the gap between group-level and individual-level effects. In mesial temporal lobe epilepsy with hippocampal sclerosis, group analyses report large reductions in ipsilateral hippocampal volume (d = 1.73-1.91, AUC ≈ 0.90).^10^ Here, hippocampal asymmetry separated unilateral mesial temporal sclerosis from non-lesional epilepsy, but marked volume loss (≥3 SD) was present in only a third of patients. Hippocampal sclerosis is diagnosed from T2/FLAIR hyperintensity and loss of internal architecture as well as atrophy, and these features can occur without measurable volume loss on T1-weighted MRI. Even for this lesion, the large effect reported in group analyses did not translate into consistent individual-level deviation.

In the primary analyses, no summary-level measure separated the groups in either within-epilepsy comparison, and differences were confined to short along-tract segments. These findings were exploratory, although the focal-generalized differences were directionally consistent with a meta-analysis reporting more pronounced white matter change in focal epilepsy.^12^ The segments were predominantly in long association and commissural pathways, which have been shown to be affected across epilepsy syndromes.^11^ Such a shared signal could produce weak group-level differences while lacking the specificity to distinguish focal from generalized or drug-resistant from drug-responsive epilepsy. Because epileptogenic networks vary across patients, the deviation common to a clinical group is likely small, suggesting that classification may require multimodal features rather than further refinement of single-modality summary scores.

Several limitations should be considered. This was a single-site, cross-sectional study, and clinical diagnosis, epilepsy type, and drug-response status were determined by retrospective review rather than independent adjudication or a uniform confirmatory protocol, so misclassification cannot be excluded. Because normative z-scores were already age- and sex-referenced, no further covariate adjustment was applied, leaving within-epilepsy comparisons unadjusted for clinical differences between groups. Epilepsy duration, for example, was longer in drug-resistant patients, possibly reflecting referral patterns, and could contribute to the diffusivity differences observed. Without presurgical evaluation, the epileptogenic zone could not be localized, and without seizure frequency data, imaging differences could not be separated from cumulative seizure burden. Excluding patients with prior brain surgery was necessary for methodological consistency but may have omitted the most severe cases.

The normative references introduced further limitations. CentileBrain model fit varied across regions (Supplementary Table 2), with the weakest fit in the medial occipital cortex (explained variance 0.08-0.14 for pericalcarine), and some composites deviated from zero in every group (e.g., occipital, thalamus, and basal ganglia; Figs. 2-4), probably reflecting site calibration. Because the same models were applied to all groups, such offsets should largely cancel in between-group comparisons, although age-dependent miscalibration could bias comparisons between groups that differ in age. CentileBrain also conditions regional cortical thickness on mean cortical thickness, so diffuse thinning would not be captured by the regional composites. The clinical diffusion protocol (single shell, 30 directions, b = 1000 s/mm², 2.2 mm) differed in angular and spatial resolution from the HCP reference acquisitions,^40^ and HCP Aging, which lacks a b = 1000 s/mm² shell, was fit at b = 1500 s/mm^2^, which increases FA and decreases MD. Because this shift coincides with the age boundary between the two HCP cohorts, residual protocol bias after ComBat harmonization^31^ could distort the normative age curve, an effect most relevant to the focal versus generalized comparison, in which groups differed in age (Supplementary Fig. 4). Finally, 30-direction diffusion imaging is not part of consensus epilepsy MRI protocols such as HARNESS-MRI^41^ and is not acquired at all centers, limiting the applicability of the tractometry analyses.

Future work could focus on the networks implicated in each patient’s epilepsy. Normative modeling yields patient-specific, spatially distributed deviations, but groups here were compared one region or tract at a time, so deviations confined to a patient’s epileptogenic network would be diluted by unaffected regions and by other patients’ differently located deviations. Localization from semiology, EEG, or presurgical evaluation could restrict analyses to the presumed epileptogenic network and align deviations to the focus. Alternatively, clustering deviation patterns across patients could identify subtypes and the modalities best suited to stratify them. Finer spatial sampling may also help, as cortical deviations here were averaged within large parcels and lobes, and the reconstructed bundles omitted superficial white matter, where abnormalities in temporal lobe epilepsy are prominent.^42,43^ Finally, the diffusion tensor assumes Gaussian diffusion and cannot capture the non-Gaussian diffusion arising from tissue microstructure; in temporal lobe epilepsy, non-Gaussian measures, which require multi-shell acquisitions, showed greater abnormality than conventional diffusivity in association white matter.^44^

In a consecutive cohort imaged on a clinical epilepsy protocol, normative modeling of structural and diffusion MRI could not classify epilepsy by type or treatment response and separated epilepsy from functional seizures only weakly, through whole-brain deviation burden. Upper confidence limits remained below the discrimination required for clinical use, indicating that any individual-level signal in univariate deviation summaries is too small to be clinically informative. Normative modeling remains a powerful framework for characterizing distributed abnormalities in epilepsy, but regional deviation scores from clinical MRI are not yet ready to guide these classifications, and network-targeted, multimodal approaches may be needed to achieve clinically useful discrimination.

## Acknowledgment

This work was supported by the National Institutes of Health (5T32NS091008-07, 5T32NS091006-10, K99NS138680, R00NS138680, R01NS148413, R01NS116504, U24NS134536, and R33NS125568), the American Epilepsy Society (Research and Training Fellowship for Clinicians), and the Zekavat Family Fund.

