## Supplemental File for "Normative Modeling of Structural and Diffusion MRI for Epilepsy Diagnosis and Classification"

**Figures**

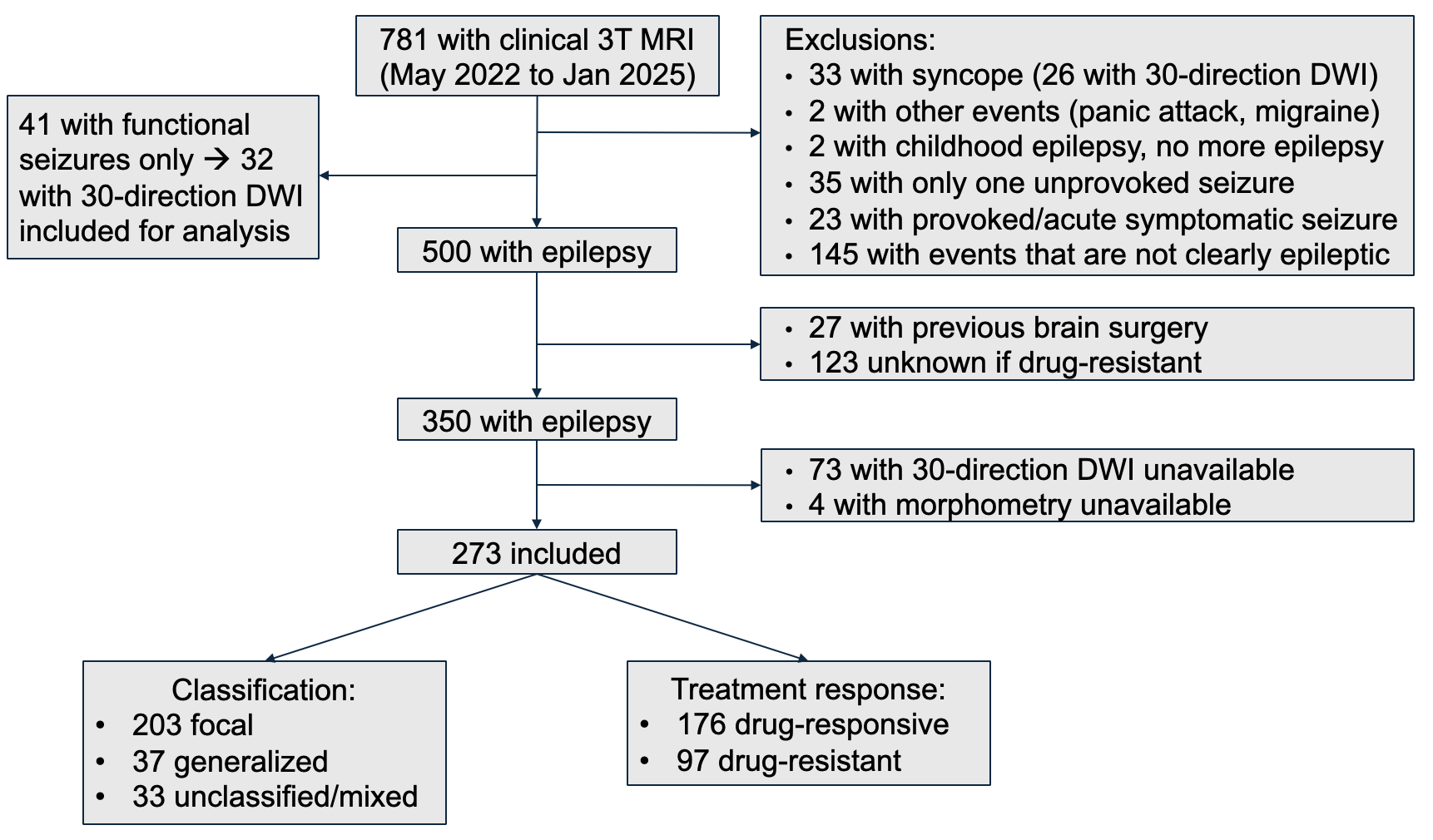

**Figure S1.** Patient selection and clinical cohort definitions. Flow of inclusion and exclusion from initial screening to the analyzed clinical cohort. Of the 33 patients with syncope, 26 with 30-direction DWI served as tractometry reference subjects. DWI, diffusion-weighted imaging.

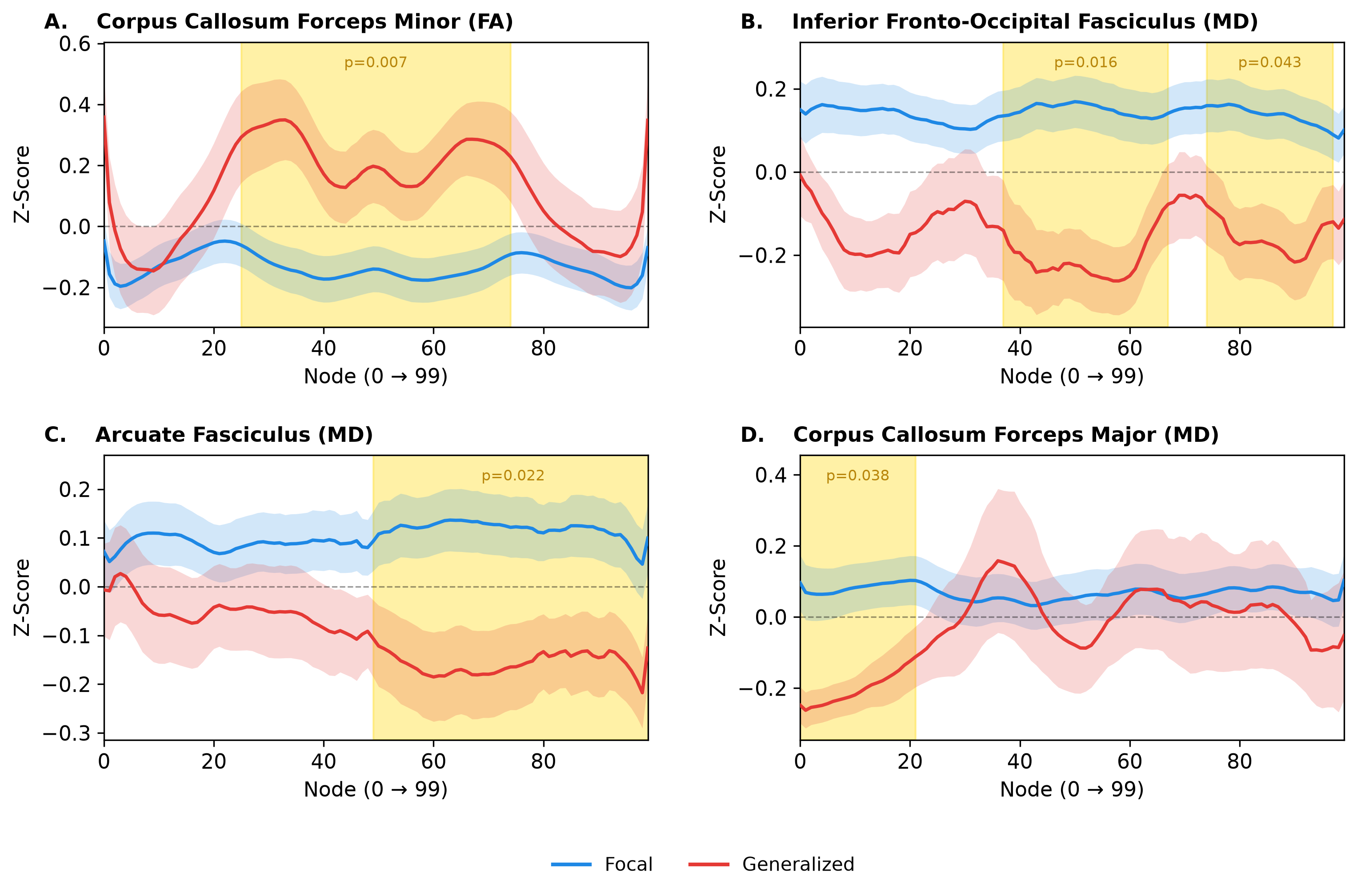

**Figure S2.** Along-tract profiles in focal vs. generalized epilepsy. Lines show mean z-scores at each of 100 nodes, and bands show ±1 SEM, for tracts with significant group differences: (A) forceps minor, fractional anisotropy (FA); (B) inferior fronto-occipital fasciculus, mean diffusivity (MD); (C) arcuate fasciculus, MD; (D) forceps major, MD. Paired tracts are averaged across hemispheres. Gold shading marks significant clusters from permutation testing (5,000 permutations), with cluster-level *p*-values; error is controlled within each tract but not across tracts. Groups are unadjusted for covariates, and all analyses are exploratory.

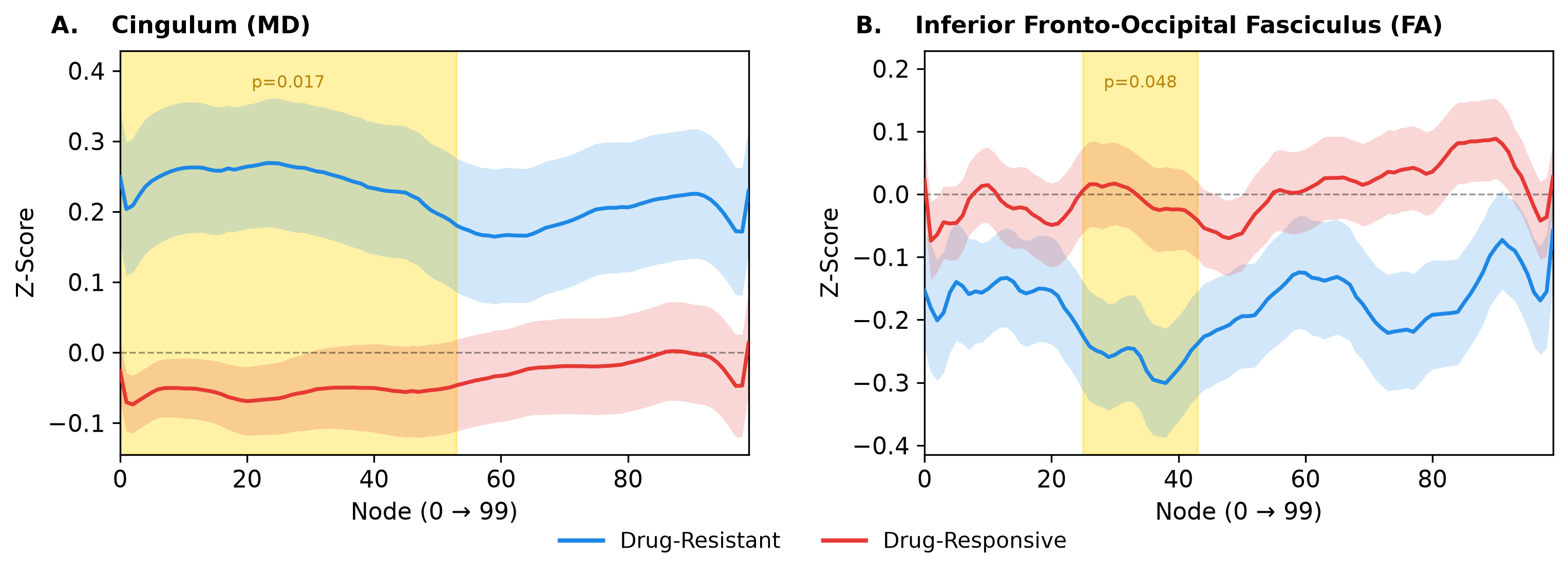

**Figure S3.** Along-tract profiles in drug-resistant vs. drug-responsive epilepsy. Lines show mean z-scores at each of 100 nodes, and bands show ±1 SEM, for tracts with significant group differences: (A) cingulum, mean diffusivity (MD); (B) inferior fronto-occipital fasciculus, fractional anisotropy (FA). Paired tracts are averaged across hemispheres. Gold shading marks significant clusters from permutation testing (5,000 permutations), with cluster-level *p*-values; error is controlled within each tract but not across tracts. Groups are unadjusted for covariates, and all analyses are exploratory.

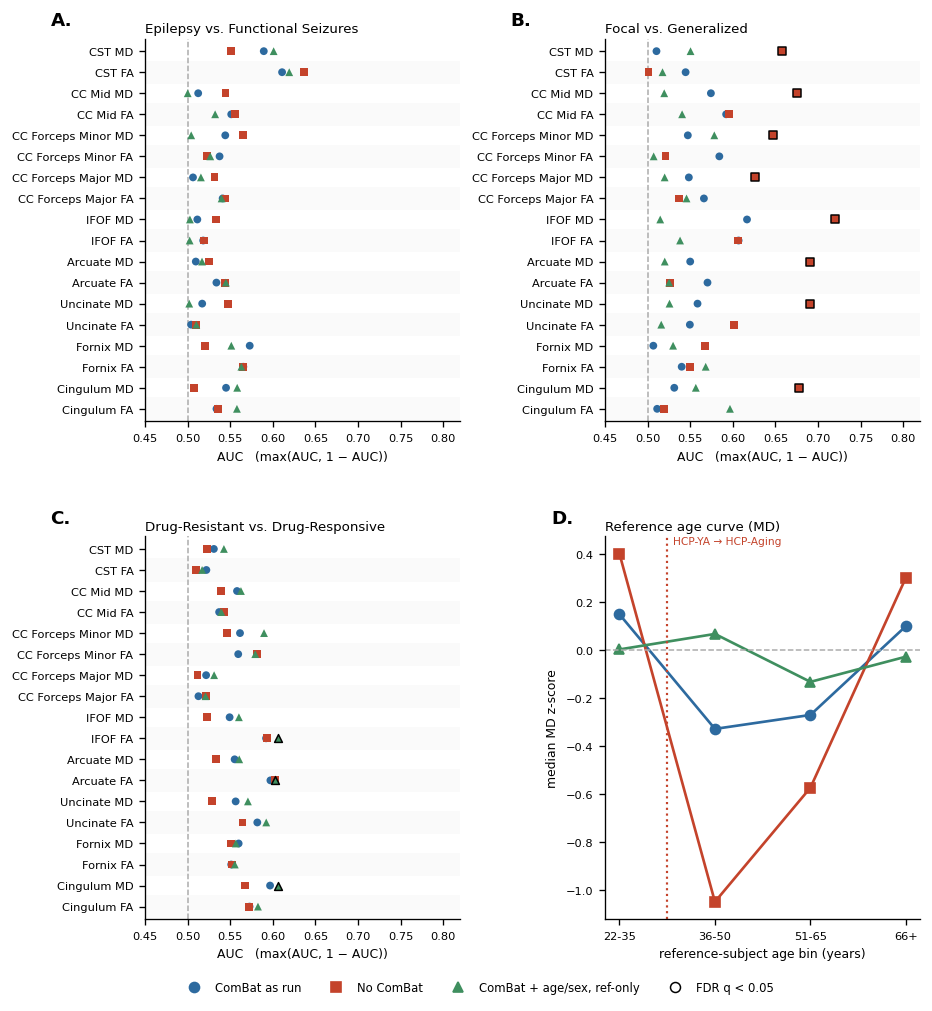

**Figure S4.** Tractometry discrimination under three harmonization schemes. (A-C) Whole-tract discrimination, shown as max(AUC, 1 − AUC), for each tract and metric in the three comparisons, with ComBat as run (circles), without ComBat (squares), and with ComBat site parameters estimated from reference subjects only (triangles); outlined markers indicate FDR-corrected *q*<0.05. (D) Median MD z-score of reference subjects by age bin under each scheme; the dotted line marks the transition from HCP Young Adult to HCP Aging. The fornix here is represented by the right fornix only, because missing left-fornix values were imputed in the main analysis; fornix AUCs therefore differ slightly from Supplementary Table 1.

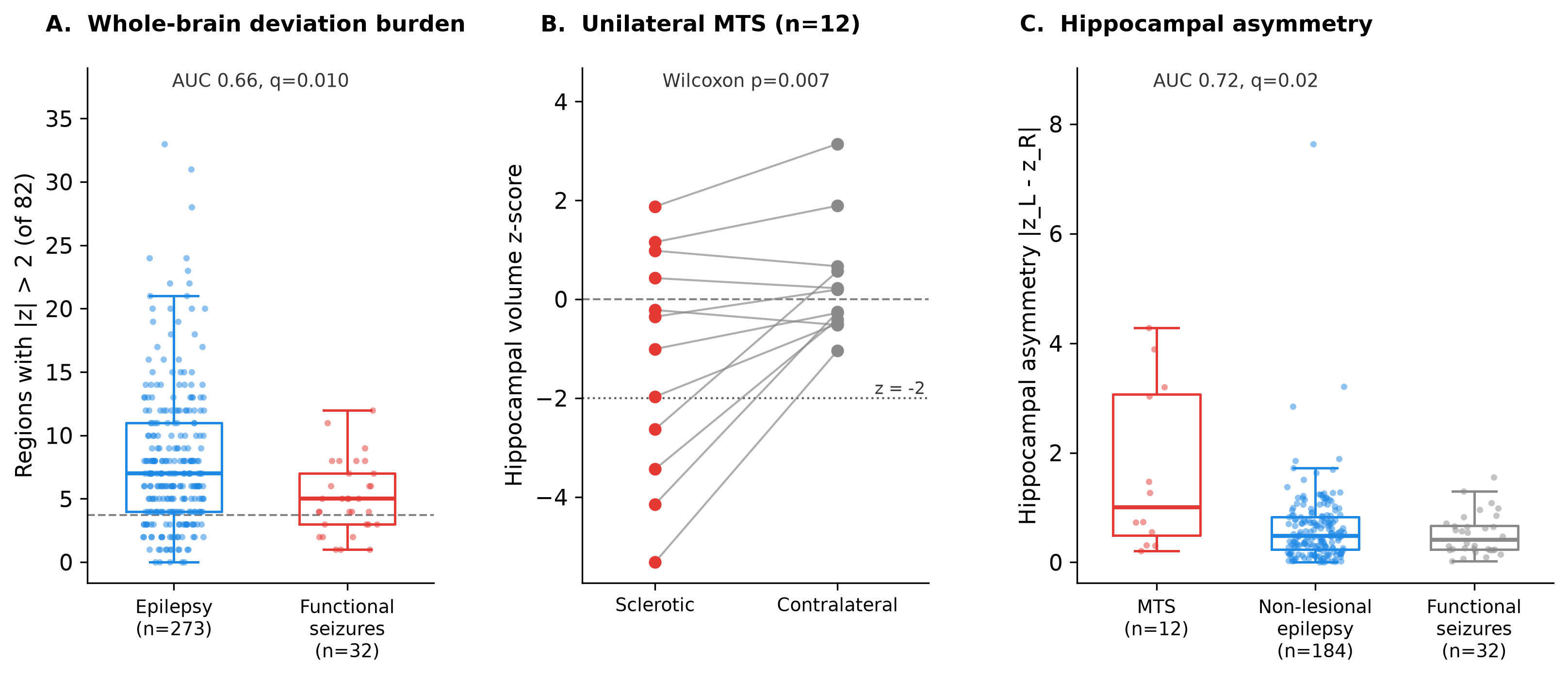

**Figure S5.** Whole-brain deviation burden and hippocampal positive control. (A) Number of the 82 morphometric regions with |*z*|>2 per patient in epilepsy and functional seizures (AUC 0.66, *q*=0.010); the dashed line marks the number expected by chance in a healthy population (4.6% of 82). (B) Hippocampal volume z-scores of the sclerotic and contralateral hippocampus in the 12 patients with unilateral mesial temporal sclerosis (MTS) and no other lesion (Wilcoxon p=0.007); the dotted line marks *z*=−2. (C) Hippocampal asymmetry (|*z*_left − *z*_right|) in MTS, non-lesional epilepsy, and functional seizures; MTS vs non-lesional epilepsy AUC 0.72, *q*=0.02. Boxes show median and IQR; points are individual patients.

**Tables**

**Table S1. Discrimination of all regional, tract-level, whole-brain, and hippocampal features**

| **Measure** | **Feature** | **AUC (95% CI)** | **Higher in** | **p** | **q** |
| --- | --- | --- | --- | --- | --- |
| **Epilepsy vs. functional seizures (n=273 vs. 32)** | | | | | |
| Lobar composite | Frontal | 0.57 (0.48-0.65) | Epilepsy | 0.200 | 0.698 |
|  | Temporal | 0.55 (0.46-0.65) | Epilepsy | 0.316 | 0.737 |
|  | Parietal | 0.59 (0.50-0.69) | Functional seizures | 0.081 | 0.570 |
|  | Occipital | 0.53 (0.43-0.63) | Epilepsy | 0.622 | 0.845 |
|  | Insula | 0.53 (0.43-0.63) | Functional seizures | 0.594 | 0.845 |
|  | Thalamus | 0.50 (0.40-0.60) | Epilepsy | 0.989 | 0.989 |
|  | Basal ganglia | 0.52 (0.43-0.61) | Functional seizures | 0.724 | 0.845 |
| Hemispheric asymmetry | Frontal | 0.57 (0.47-0.68) | Functional seizures | 0.175 | 0.307 |
|  | Temporal | 0.60 (0.51-0.69) | Epilepsy | 0.056 | 0.197 |
|  | Parietal | 0.55 (0.45-0.65) | Epilepsy | 0.376 | 0.439 |
|  | Occipital | 0.61 (0.50-0.72) | Functional seizures | 0.038 | 0.197 |
|  | Insula | 0.55 (0.45-0.65) | Epilepsy | 0.370 | 0.439 |
|  | Thalamus | 0.53 (0.41-0.65) | Epilepsy | 0.569 | 0.569 |
|  | Basal ganglia | 0.59 (0.49-0.69) | Epilepsy | 0.102 | 0.237 |
| Tract-level FA | Cingulum | 0.53 (0.42-0.64) | Epilepsy | 0.531 | 0.872 |
|  | Fornix | 0.56 (0.45-0.66) | Epilepsy | 0.304 | 0.872 |
|  | Uncinate fasciculus | 0.50 (0.40-0.61) | Functional seizures | 0.940 | 0.940 |
|  | Arcuate fasciculus | 0.53 (0.42-0.64) | Epilepsy | 0.533 | 0.872 |
|  | Inferior fronto-occipital fasciculus | 0.52 (0.41-0.63) | Functional seizures | 0.735 | 0.940 |
|  | Corpus callosum, forceps major | 0.54 (0.43-0.65) | Epilepsy | 0.448 | 0.872 |
|  | Corpus callosum, forceps minor | 0.54 (0.43-0.65) | Functional seizures | 0.488 | 0.872 |
|  | Corpus callosum, body | 0.55 (0.44-0.66) | Functional seizures | 0.344 | 0.872 |
|  | Corticospinal tract | 0.61 (0.50-0.72) | Epilepsy | 0.040 | 0.723 |
| Tract-level MD | Cingulum | 0.55 (0.43-0.65) | Functional seizures | 0.404 | 0.872 |
|  | Fornix | 0.54 (0.43-0.64) | Functional seizures | 0.520 | 0.872 |
|  | Uncinate fasciculus | 0.52 (0.42-0.62) | Epilepsy | 0.753 | 0.940 |
|  | Arcuate fasciculus | 0.51 (0.40-0.62) | Functional seizures | 0.860 | 0.940 |
|  | Inferior fronto-occipital fasciculus | 0.51 (0.41-0.62) | Epilepsy | 0.833 | 0.940 |
|  | Corpus callosum, forceps major | 0.51 (0.40-0.61) | Epilepsy | 0.908 | 0.940 |
|  | Corpus callosum, forceps minor | 0.54 (0.44-0.65) | Epilepsy | 0.414 | 0.872 |
|  | Corpus callosum, body | 0.51 (0.41-0.62) | Epilepsy | 0.820 | 0.940 |
|  | Corticospinal tract | 0.59 (0.49-0.69) | Functional seizures | 0.098 | 0.872 |
| Whole-brain burden | Morphometry (82), count \|z\|>2 | 0.66 (0.57-0.74) | Epilepsy | 0.003 | 0.009 |
|  | Morphometry (82), mean \|z\| | 0.67 (0.58-0.76) | Epilepsy | 0.002 | 0.009 |
|  | Tract-level (18), count \|z\|>2 | 0.54 (0.50-0.57) | Epilepsy | 0.154 | 0.232 |
|  | Tract-level (18), mean \|z\| | 0.51 (0.40-0.61) | Functional seizures | 0.933 | 0.933 |
|  | Node-level (3,000), count \|z\|>2 | 0.60 (0.49-0.71) | Epilepsy | 0.056 | 0.113 |
|  | Node-level (3,000), mean \|z\| | 0.54 (0.43-0.65) | Epilepsy | 0.432 | 0.519 |
| Hippocampal volume | Asymmetry index | 0.57 (0.48-0.66) | Epilepsy | 0.184 | 0.369 |
|  | Mean z-score | 0.54 (0.45-0.62) | Epilepsy | 0.488 | 0.488 |
| **Focal vs. generalized epilepsy (n=203 vs. 37)** | | | | | |
| Lobar composite | Frontal | 0.54 (0.43-0.64) | Focal | 0.477 | 0.683 |
|  | Temporal | 0.55 (0.46-0.65) | Focal | 0.312 | 0.683 |
|  | Parietal | 0.50 (0.40-0.60) | Generalized | 0.994 | 0.994 |
|  | Occipital | 0.53 (0.43-0.62) | Focal | 0.566 | 0.683 |
|  | Insula | 0.53 (0.43-0.63) | Focal | 0.585 | 0.683 |
|  | Thalamus | 0.53 (0.43-0.63) | Focal | 0.571 | 0.683 |
|  | Basal ganglia | 0.54 (0.44-0.64) | Focal | 0.479 | 0.683 |
| Hemispheric asymmetry | Frontal | 0.55 (0.45-0.64) | Focal | 0.383 | 0.447 |
|  | Temporal | 0.56 (0.45-0.66) | Focal | 0.261 | 0.447 |
|  | Parietal | 0.55 (0.45-0.65) | Focal | 0.338 | 0.447 |
|  | Occipital | 0.50 (0.41-0.60) | Generalized | 0.953 | 0.953 |
|  | Insula | 0.59 (0.49-0.69) | Generalized | 0.086 | 0.353 |
|  | Thalamus | 0.58 (0.49-0.67) | Focal | 0.101 | 0.353 |
|  | Basal ganglia | 0.57 (0.47-0.67) | Focal | 0.183 | 0.427 |
| Tract-level FA | Cingulum | 0.51 (0.41-0.61) | Focal | 0.831 | 0.843 |
|  | Fornix | 0.55 (0.45-0.65) | Focal | 0.329 | 0.501 |
|  | Uncinate fasciculus | 0.55 (0.45-0.64) | Generalized | 0.340 | 0.501 |
|  | Arcuate fasciculus | 0.57 (0.47-0.67) | Generalized | 0.175 | 0.501 |
|  | Inferior fronto-occipital fasciculus | 0.61 (0.51-0.70) | Generalized | 0.039 | 0.353 |
|  | Corpus callosum, forceps major | 0.57 (0.46-0.66) | Generalized | 0.203 | 0.501 |
|  | Corpus callosum, forceps minor | 0.58 (0.49-0.68) | Generalized | 0.104 | 0.469 |
|  | Corpus callosum, body | 0.59 (0.48-0.69) | Generalized | 0.075 | 0.451 |
|  | Corticospinal tract | 0.55 (0.45-0.64) | Generalized | 0.390 | 0.501 |
| Tract-level MD | Cingulum | 0.53 (0.43-0.62) | Focal | 0.547 | 0.656 |
|  | Fornix | 0.52 (0.42-0.62) | Generalized | 0.686 | 0.772 |
|  | Uncinate fasciculus | 0.56 (0.46-0.65) | Focal | 0.258 | 0.501 |
|  | Arcuate fasciculus | 0.55 (0.46-0.64) | Focal | 0.336 | 0.501 |
|  | Inferior fronto-occipital fasciculus | 0.62 (0.53-0.70) | Focal | 0.024 | 0.353 |
|  | Corpus callosum, forceps major | 0.55 (0.45-0.64) | Focal | 0.351 | 0.501 |
|  | Corpus callosum, forceps minor | 0.55 (0.45-0.64) | Focal | 0.363 | 0.501 |
|  | Corpus callosum, body | 0.57 (0.48-0.67) | Focal | 0.151 | 0.501 |
|  | Corticospinal tract | 0.51 (0.41-0.61) | Focal | 0.843 | 0.843 |
| Whole-brain burden | Morphometry (82), count \|z\|>2 | 0.52 (0.43-0.62) | Generalized | 0.673 | 0.715 |
|  | Morphometry (82), mean \|z\| | 0.52 (0.42-0.61) | Generalized | 0.715 | 0.715 |
|  | Tract-level (18), count \|z\|>2 | 0.53 (0.49-0.57) | Focal | 0.229 | 0.459 |
|  | Tract-level (18), mean \|z\| | 0.55 (0.44-0.64) | Focal | 0.387 | 0.581 |
|  | Node-level (3,000), count \|z\|>2 | 0.56 (0.46-0.66) | Focal | 0.230 | 0.459 |
|  | Node-level (3,000), mean \|z\| | 0.58 (0.48-0.68) | Focal | 0.116 | 0.459 |
| Hippocampal volume | Asymmetry index | 0.57 (0.48-0.67) | Focal | 0.154 | 0.154 |
|  | Mean z-score | 0.59 (0.49-0.68) | Focal | 0.083 | 0.154 |
| **Drug-resistant vs. drug-responsive epilepsy (n=97 vs. 176)** | | | | | |
| Lobar composite | Frontal | 0.52 (0.45-0.59) | Drug-responsive | 0.578 | 0.722 |
|  | Temporal | 0.52 (0.45-0.58) | Drug-responsive | 0.665 | 0.722 |
|  | Parietal | 0.53 (0.45-0.60) | Drug-responsive | 0.491 | 0.722 |
|  | Occipital | 0.51 (0.44-0.58) | Drug-resistant | 0.722 | 0.722 |
|  | Insula | 0.53 (0.46-0.60) | Drug-resistant | 0.429 | 0.722 |
|  | Thalamus | 0.57 (0.49-0.64) | Drug-responsive | 0.070 | 0.246 |
|  | Basal ganglia | 0.58 (0.51-0.65) | Drug-responsive | 0.031 | 0.217 |
| Hemispheric asymmetry | Frontal | 0.56 (0.49-0.64) | Drug-resistant | 0.077 | 0.270 |
|  | Temporal | 0.52 (0.45-0.59) | Drug-responsive | 0.528 | 0.740 |
|  | Parietal | 0.57 (0.50-0.65) | Drug-responsive | 0.050 | 0.270 |
|  | Occipital | 0.51 (0.43-0.58) | Drug-responsive | 0.883 | 1.000 |
|  | Insula | 0.55 (0.48-0.62) | Drug-resistant | 0.209 | 0.487 |
|  | Thalamus | 0.53 (0.46-0.60) | Drug-resistant | 0.411 | 0.719 |
|  | Basal ganglia | 0.50 (0.43-0.57) | Drug-resistant | 1.000 | 1.000 |
| Tract-level FA | Cingulum | 0.57 (0.50-0.65) | Drug-responsive | 0.047 | 0.169 |
|  | Fornix | 0.50 (0.43-0.57) | Drug-responsive | 0.941 | 0.941 |
|  | Uncinate fasciculus | 0.58 (0.51-0.65) | Drug-responsive | 0.025 | 0.114 |
|  | Arcuate fasciculus | 0.60 (0.53-0.67) | Drug-responsive | 0.008 | 0.071 |
|  | Inferior fronto-occipital fasciculus | 0.59 (0.52-0.66) | Drug-responsive | 0.012 | 0.071 |
|  | Corpus callosum, forceps major | 0.51 (0.44-0.59) | Drug-responsive | 0.729 | 0.772 |
|  | Corpus callosum, forceps minor | 0.56 (0.49-0.63) | Drug-responsive | 0.104 | 0.237 |
|  | Corpus callosum, body | 0.54 (0.46-0.61) | Drug-responsive | 0.313 | 0.433 |
|  | Corticospinal tract | 0.52 (0.45-0.60) | Drug-resistant | 0.548 | 0.623 |
| Tract-level MD | Cingulum | 0.60 (0.52-0.67) | Drug-resistant | 0.008 | 0.071 |
|  | Fornix | 0.54 (0.47-0.62) | Drug-resistant | 0.246 | 0.370 |
|  | Uncinate fasciculus | 0.56 (0.48-0.63) | Drug-resistant | 0.123 | 0.237 |
|  | Arcuate fasciculus | 0.56 (0.48-0.63) | Drug-resistant | 0.132 | 0.237 |
|  | Inferior fronto-occipital fasciculus | 0.55 (0.48-0.62) | Drug-resistant | 0.178 | 0.291 |
|  | Corpus callosum, forceps major | 0.52 (0.45-0.59) | Drug-resistant | 0.554 | 0.623 |
|  | Corpus callosum, forceps minor | 0.56 (0.49-0.63) | Drug-resistant | 0.093 | 0.237 |
|  | Corpus callosum, body | 0.56 (0.49-0.63) | Drug-resistant | 0.113 | 0.237 |
|  | Corticospinal tract | 0.53 (0.46-0.60) | Drug-resistant | 0.401 | 0.515 |
| Whole-brain burden | Morphometry (82), count \|z\|>2 | 0.51 (0.44-0.58) | Drug-resistant | 0.817 | 0.817 |
|  | Morphometry (82), mean \|z\| | 0.51 (0.44-0.58) | Drug-resistant | 0.781 | 0.817 |
|  | Tract-level (18), count \|z\|>2 | 0.51 (0.47-0.55) | Drug-resistant | 0.738 | 0.817 |
|  | Tract-level (18), mean \|z\| | 0.52 (0.45-0.59) | Drug-resistant | 0.549 | 0.817 |
|  | Node-level (3,000), count \|z\|>2 | 0.55 (0.48-0.63) | Drug-resistant | 0.152 | 0.817 |
|  | Node-level (3,000), mean \|z\| | 0.54 (0.47-0.61) | Drug-resistant | 0.285 | 0.817 |
| Hippocampal volume | Asymmetry index | 0.54 (0.47-0.61) | Drug-resistant | 0.269 | 0.279 |
|  | Mean z-score | 0.54 (0.47-0.61) | Drug-responsive | 0.279 | 0.279 |
| **Unilateral MTS vs. non-lesional epilepsy (n=12 vs. 184)** | | | | | |
| Hippocampal volume | Asymmetry index | 0.72 (0.55-0.88) | MTS | 0.012 | 0.023 |
|  | Mean z-score | 0.63 (0.43-0.81) | Non-lesional epilepsy | 0.139 | 0.139 |
| **Non-lesional epilepsy vs. functional seizures (n=184 vs. 32)** | | | | | |
| Whole-brain burden | Morphometry (82), count \|z\|>2 | 0.62 (0.53-0.72) | Non-lesional epilepsy | 0.026 | 0.080 |
|  | Morphometry (82), mean \|z\| | 0.64 (0.54-0.74) | Non-lesional epilepsy | 0.011 | 0.067 |
|  | Tract-level (18), count \|z\|>2 | 0.52 (0.48-0.55) | Non-lesional epilepsy | 0.379 | 0.568 |
|  | Tract-level (18), mean \|z\| | 0.53 (0.42-0.64) | Functional seizures | 0.582 | 0.699 |
|  | Node-level (3,000), count \|z\|>2 | 0.56 (0.44-0.68) | Non-lesional epilepsy | 0.256 | 0.512 |
|  | Node-level (3,000), mean \|z\| | 0.51 (0.39-0.62) | Non-lesional epilepsy | 0.930 | 0.930 |

**Abbreviations:** AUC, area under the receiver operating characteristic curve; CI, confidence interval; FA, fractional anisotropy; FDR, false discovery rate; MD, mean diffusivity; MTS, mesial temporal sclerosis; z_L and z_R, left and right hemisphere z-scores.

**Table S2. CentileBrain Normative Model Fit Statistics**

| Hemisphere | Region | Corr (Male) | EV (Male) | Corr (Female) | EV (Female) |
| --- | --- | --- | --- | --- | --- |
| Left | Banks of superior temporal sulcus | 0.67 | 0.44 | 0.68 | 0.44 |
| Left | Caudal anterior cingulate | 0.55 | 0.28 | 0.42 | 0.18 |
| Left | Caudal middle frontal | 0.86 | 0.74 | 0.78 | 0.61 |
| Left | Cuneus | 0.58 | 0.33 | 0.51 | 0.25 |
| Left | Entorhinal | 0.39 | 0.15 | 0.36 | 0.12 |
| Left | Fusiform | 0.8 | 0.61 | 0.8 | 0.6 |
| Left | Inferior parietal | 0.89 | 0.79 | 0.8 | 0.65 |
| Left | Inferior temporal | 0.77 | 0.59 | 0.73 | 0.53 |
| Left | Isthmus cingulate | 0.61 | 0.37 | 0.44 | 0.19 |
| Left | Lateral occipital | 0.77 | 0.59 | 0.7 | 0.49 |
| Left | Lateral orbitofrontal | 0.77 | 0.59 | 0.56 | 0.3 |
| Left | Lingual | 0.62 | 0.39 | 0.57 | 0.32 |
| Left | Medial orbitofrontal | 0.74 | 0.51 | 0.42 | 0.17 |
| Left | Middle temporal | 0.8 | 0.64 | 0.73 | 0.53 |
| Left | Parahippocampal | 0.44 | 0.19 | 0.39 | 0.15 |
| Left | Paracentral | 0.8 | 0.64 | 0.79 | 0.6 |
| Left | Pars opercularis | 0.8 | 0.64 | 0.75 | 0.56 |
| Left | Pars orbitalis | 0.6 | 0.35 | 0.4 | 0.11 |
| Left | Pars triangularis | 0.74 | 0.54 | 0.62 | 0.37 |
| Left | Pericalcarine | 0.4 | 0.14 | 0.31 | 0.08 |
| Left | Postcentral | 0.76 | 0.57 | 0.63 | 0.38 |
| Left | Posterior cingulate | 0.68 | 0.46 | 0.62 | 0.38 |
| Left | Precentral | 0.77 | 0.6 | 0.74 | 0.55 |
| Left | Precuneus | 0.89 | 0.78 | 0.81 | 0.65 |
| Left | Rostral anterior cingulate | 0.62 | 0.39 | 0.42 | 0.17 |
| Left | Rostral middle frontal | 0.83 | 0.69 | 0.7 | 0.47 |
| Left | Superior frontal | 0.9 | 0.8 | 0.83 | 0.69 |
| Left | Superior parietal | 0.87 | 0.75 | 0.83 | 0.68 |
| Left | Superior temporal | 0.85 | 0.72 | 0.77 | 0.59 |
| Left | Supramarginal | 0.87 | 0.76 | 0.8 | 0.64 |
| Left | Frontal pole | 0.58 | 0.33 | 0.31 | 0.09 |
| Left | Temporal pole | 0.4 | 0.16 | 0.19 | 0.01 |
| Left | Transverse temporal | 0.57 | 0.33 | 0.58 | 0.34 |
| Left | Insula | 0.69 | 0.48 | 0.67 | 0.44 |
| Left | Thalamus | 0.67 | 0.41 | 0.76 | 0.52 |
| Left | Caudate | 0.5 | 0.25 | 0.6 | 0.36 |
| Left | Putamen | 0.57 | 0.32 | 0.57 | 0.31 |
| Left | Pallidum | 0.54 | 0.28 | 0.67 | 0.42 |
| Left | Hippocampus | 0.5 | 0.24 | 0.75 | 0.48 |
| Left | Amygdala | 0.4 | 0.16 | 0.61 | 0.35 |
| Left | Nucleus accumbens | 0.52 | 0.27 | 0.52 | 0.27 |
| Right | Banks of superior temporal sulcus | 0.7 | 0.49 | 0.74 | 0.5 |
| Right | Caudal anterior cingulate | 0.58 | 0.31 | 0.47 | 0.22 |
| Right | Caudal middle frontal | 0.74 | 0.53 | 0.83 | 0.67 |
| Right | Cuneus | 0.63 | 0.4 | 0.53 | 0.28 |
| Right | Entorhinal | 0.39 | 0.15 | 0.28 | 0.08 |
| Right | Fusiform | 0.73 | 0.54 | 0.73 | 0.52 |
| Right | Inferior parietal | 0.9 | 0.81 | 0.83 | 0.67 |
| Right | Inferior temporal | 0.73 | 0.54 | 0.7 | 0.48 |
| Right | Isthmus cingulate | 0.57 | 0.33 | 0.43 | 0.18 |
| Right | Lateral occipital | 0.76 | 0.57 | 0.72 | 0.51 |
| Right | Lateral orbitofrontal | 0.76 | 0.57 | 0.54 | 0.29 |
| Right | Lingual | 0.56 | 0.29 | 0.56 | 0.31 |
| Right | Medial orbitofrontal | 0.7 | 0.46 | 0.53 | 0.28 |
| Right | Middle temporal | 0.78 | 0.6 | 0.8 | 0.63 |
| Right | Parahippocampal | 0.44 | 0.19 | 0.43 | 0.18 |
| Right | Paracentral | 0.77 | 0.59 | 0.77 | 0.58 |
| Right | Pars opercularis | 0.75 | 0.55 | 0.76 | 0.56 |
| Right | Pars orbitalis | 0.74 | 0.54 | 0.51 | 0.23 |
| Right | Pars triangularis | 0.75 | 0.55 | 0.75 | 0.57 |
| Right | Pericalcarine | 0.35 | 0.1 | 0.35 | 0.12 |
| Right | Postcentral | 0.7 | 0.49 | 0.61 | 0.37 |
| Right | Posterior cingulate | 0.68 | 0.46 | 0.54 | 0.29 |
| Right | Precentral | 0.66 | 0.43 | 0.75 | 0.56 |
| Right | Precuneus | 0.86 | 0.73 | 0.81 | 0.66 |
| Right | Rostral anterior cingulate | 0.63 | 0.38 | 0.5 | 0.24 |
| Right | Rostral middle frontal | 0.74 | 0.55 | 0.64 | 0.38 |
| Right | Superior frontal | 0.86 | 0.74 | 0.82 | 0.67 |
| Right | Superior parietal | 0.86 | 0.74 | 0.8 | 0.64 |
| Right | Superior temporal | 0.85 | 0.72 | 0.84 | 0.69 |
| Right | Supramarginal | 0.85 | 0.72 | 0.82 | 0.64 |
| Right | Frontal pole | 0.56 | 0.3 | 0.37 | 0.14 |
| Right | Temporal pole | 0.46 | 0.21 | 0.34 | 0.11 |
| Right | Transverse temporal | 0.66 | 0.43 | 0.61 | 0.35 |
| Right | Insula | 0.73 | 0.53 | 0.74 | 0.5 |
| Right | Thalamus | 0.67 | 0.42 | 0.71 | 0.45 |
| Right | Caudate | 0.51 | 0.26 | 0.63 | 0.4 |
| Right | Putamen | 0.57 | 0.32 | 0.59 | 0.34 |
| Right | Pallidum | 0.58 | 0.3 | 0.62 | 0.35 |
| Right | Hippocampus | 0.55 | 0.28 | 0.73 | 0.44 |
| Right | Amygdala | 0.45 | 0.2 | 0.56 | 0.3 |
| Right | Nucleus accumbens | 0.49 | 0.24 | 0.61 | 0.37 |

**Abbreviations:** Corr, correlation between predicted and observed values, with predictions based on age, sex, and intracranial volume (subcortical volumes) or mean cortical thickness (cortical thickness); EV, explained variance. Both statistics are as reported by CentileBrain for this cohort, for each region in the male and female reference models.
